# Center-Based Experiences Implementing Strategies to Reduce Risk of Horizontal Transmission of SARS-Cov-2: Potential for Compromise of Neonatal Microbiome Assemblage

**DOI:** 10.1101/2021.01.07.21249418

**Authors:** Joann Romano-Keeler, Dana Fiszbein, Jilei Zhang, Joseph Horowitz, Karen Hayani, Irina Buhimschi, Christina Lopez, Zaynab Kadhem, James Berman, Phornphat Rasamimari, Aarti Raghavan, De-Ann M. Pillers, Jun Sun

## Abstract

Perinatal transmission of COVID-19 is poorly understood and many neonatal intensive care units’ (NICU) policies minimize mother-infant contact to prevent transmission. We present our unit’s approach and ways it may impact neonatal microbiome acquisition. We attended COVID-19 positive mothers’ deliveries from March-August 2020. Delayed cord clamping and skin-to-skin were avoided and infants were admitted to the NICU. No parents’ visits were allowed and discharge was arranged with COVID-19 negative family members. Maternal breast milk was restricted in the NICU. All twenty-one infants tested negative at 24 and 48 hours and had average hospital stays of nine days. 40% of mothers expressed breastmilk and 60% of infants were discharged with COVID-19 negative caregivers. Extended hospital stays, no skin-to-skin contact, limited maternal milk use, and discharge to caregivers outside primary residences, potentially affect the neonatal microbiome. Future studies are warranted to explore how ours and other centers’ similar policies influence this outcome.

## Introduction

Since its presentation in Wuhan, China, coronavirus disease of 2019 (COVID-19) caused by severe acute respiratory syndrome coronavirus 2 (SARS-CoV-2) has evolved into a global pandemic with significant mortality and morbidity. The number of confirmed cases has climbed to over thirty million with one million deaths in more than 200 countries and territories.(1, 2) While initially associated with fever, cough and shortness of breath, presentation of this virus has progressed to include gastrointestinal (GI) symptoms either in isolation or in conjunction with respiratory symptoms in 10-15% of COVID-19 positive patients.(3, 4) GI symptoms may herald COVID-19 by up to 5 days before admission and these patients may have more complicated courses with longer hospital stays.(5, 6)

Neonatal intensive care units (NICU) and newborn nurseries have developed strategies to minimize risk of infection to the relatively immunocompromised newborn population born to COVID-19 positive mothers. American Academy of Pediatrics (AAP) recommendations addressed this issue based on best available evidence at that time, experiences from other institutions, and analysis of the risk/benefit to the mother and child.(7) Our guidelines were based on limited evidence available at the early stages of the pandemic about neonatal COVID-19 infections and our concerns about transmission in an open bay unit with limited space to socially distance.(8, 9) Our feeding polices reflected our not having a dedicated milk preparation room or a barcoded milk storage system. No existing recommendations address potential impact that perinatal management may have on early life microbial colonization and subsequent newborn and childhood health outcomes.(10, 11)

Intrinsic and extrinsic factors drive early life assembly of the intestinal microbiome, including maternal genetics and nutrition during pregnancy.(12, 13) Postnatal variables including infants’ diet, antibiotic use, and environmental exposures can shape bacterial colonization until “adult-like” gut microbiome is established.(14, 15) Virome-microbiome crosstalk during this period is poorly understood.(16) No studies have investigated interactions between COVID-19 and the nascent microbiome or how policies to prevent perinatal COVID transmission may affect newborn microbial colonization **(Figure 1)**.

**Figure 1:**
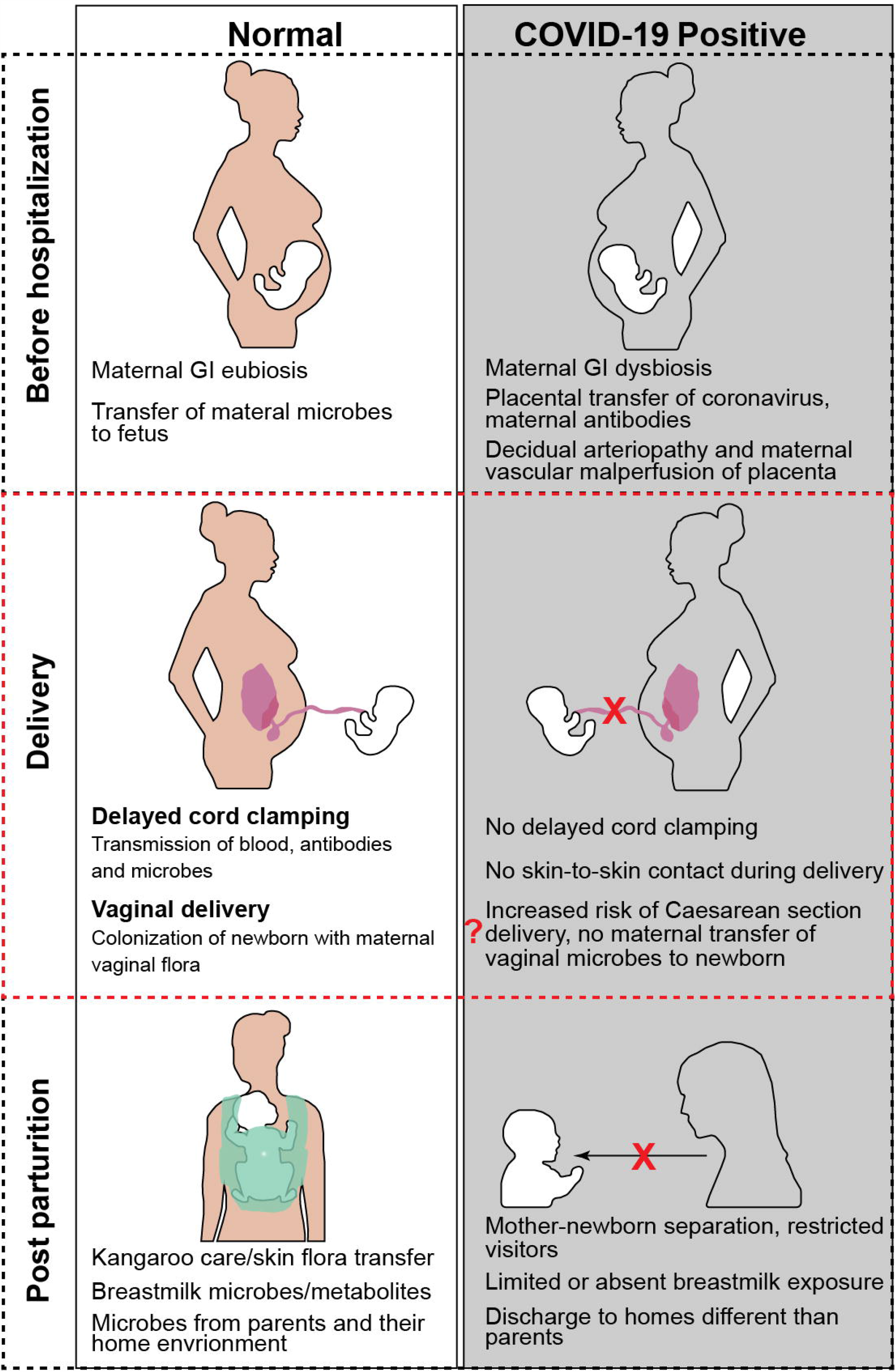
Potential interruptions in early life microbial colonization due to COVID-19 and infection control policies. Placental exchange of the maternal microbiome with the fetus during pregnancy contributes to early bacterial colonization. In contrast, placentas of women with COVID-19 have increased rates of decidual arteriopathy and maternal vascular malperfusion which may affect normal bacterial exchange.(33) At delivery, mothers with critical illness or fetal distress secondary to COVID-19 may be more likely to deliver via Cesarean section depriving the newborn of normal bacterial colonization. Institutional policies to avoid delayed cord clamping in deliveries of COVID-19 mothers may also impact microbial transfer to the newborn. Another infection control measure restricts skin-to-skin care between COVID-19 positive mothers and their infants which interferes with newborn’s inheritance of maternal oral and skin flora. In addition, we recommended discharge of the newborn to COVID negative family members if possible until COVID-19 parents complete 10 days of separation, which often resulted in discharge home with family members outside the traditional household.

Twenty-one patients born to COVID-19 positive mothers were admitted to the NICU per University of Illinois at Chicago (UIC) guidelines. In this paper, we review NICU courses, including the incidence viral transmission. We explore ways that our guidelines, designed to reduce risk of viral transmission to infants, may impact composition of the newborn microbiome. Finally, we discuss future directions for NICU management that minimize perinatal viral transmission, promote normal early life microbiome assemblage, and optimize short and long-term health outcomes.

## Materials/Subjects and Methods

This study was approved by the UIC Institutional Review Board (protocol no. 2020-0443). Neonatal teams attended deliveries of all COVID-19 positive mothers from March-August 2020. Deidentified maternal data collected included age, reported race, dates and types of COVID-19 testing, COVID-related symptoms at delivery, and details on peripartum courses including intrapartum antibiotics and timing of rupture of membranes (**Table 1**).

**Table 1.**
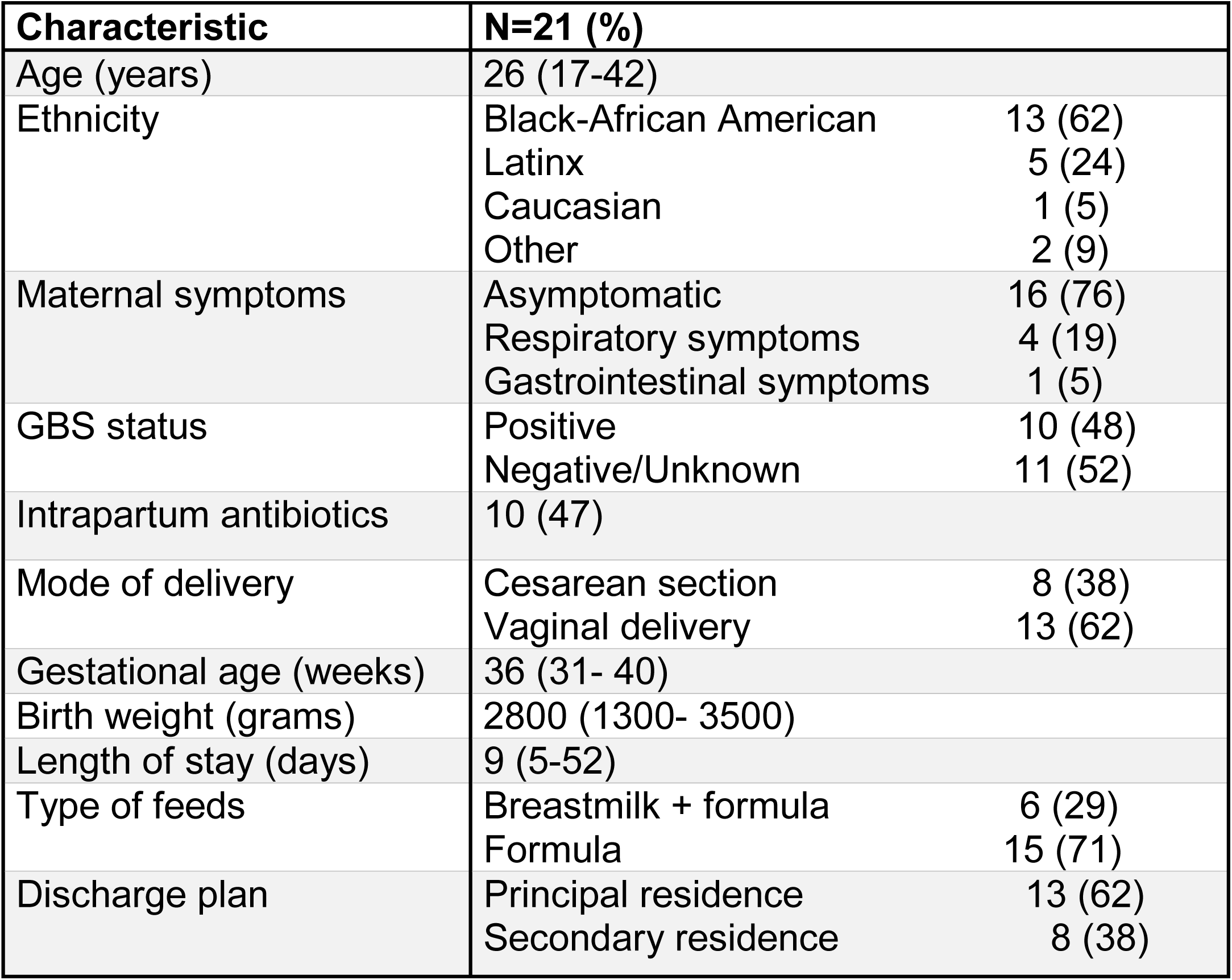
Clinical Characteristics of COVID positive mother-infant dyads.

Delayed cord clamping and skin-to-skin contact were avoided, including physical contact with family members and infants were admitted to the NICU with contact isolation. Metadata obtained on infants included delivery indication and mode, Apgar scores, gestational age, birth weight, and gender. Respiratory and/or gastrointestinal symptoms were recorded throughout hospitalization and POCT was completed at 24 and 48 hours of life. Visitors were not allowed during NICU stays. Discharges were arranged whenever possible with COVID-19 negative caregivers. No expressed breastmilk was used in the NICU but mothers were encouraged to pump and store milk and instructed on how to safely breastfeed at home. Information on feeding regimen after discharge was recorded when available from outpatient clinic notes.

## Results

### Subjects

Twenty-one COVID-19 positive mothers positive delivered during this study with a mean (range) age of 26 (17-42) years. Approximately 62% of mothers were African American (n=13) and 5% were Latinx (n=5). COVID-19 antigen detection via rapid point of care testing (POCT) within 72 hours of delivery or as an inpatient in labor and delivery was positive in 90% of mothers (n=19). The remaining two mothers had positive viral PCR tests collected prior to delivery.

### Clinical Characteristics of COVID-19 mothers

Sixteen COVID-19 positive mothers (76%) were asymptomatic and five (24%) were symptomatic, including one with gastrointestinal symptoms. Group B Streptococcus (GBS) screening was positive in 48% of mothers (n=10) and prolonged rupture of membranes (>18 hours prior to delivery) complicated one delivery. Intrapartum antibiotics were administered in 57% (n=11) of mothers, including five who were GBS positive and seven who received cefazolin perioperatively. In 38% of mothers (n=8) Caesarean sections (C-section) were performed for fetal heart rate decelerations or arrest of dilation (n=5), breech presentation (n=1), repeat C-sections (n=1), or COVID-19 related maternal morbidities (n=1).

### Infant demographics and Hospital Course

Mean [range] gestational age and birth weight was 36 [31-40] weeks and 2800 [1300-3500] grams, respectively. Prematurity complicated two deliveries, including one case where an infant was delivered early due to the severity of mother’s COVID-19 infection. In 40% of cases (n=8), delayed cord clamping occurred and in 10% (n=2) of deliveries, mothers were allowed skin-to-skin contact. Approximately 70% of infants were male with a mean one minute Apgar score of 7 and a 5 minute score of 9. POCT was negative at 24 and 48 hours for all infants. In addition, two infants had a third POCT due to persistent oxygen requirements, which were also negative. More than half of patients (n=11) required respiratory support including mechanical ventilation (n=1), continuous positive airway pressure (n=8), and nasal cannula (n=2). At the time of discharge, one patient remained on oxygen.

### Breast milk usage

Lactation counselors met with mothers to instruct on safe techniques for nursing and pumping, including pumping during NICU stays and either discarding or storing milk. All infants received formula in the NICU, except for one premature infant who qualified for donor breastmilk. In 30% of newborns, mothers provided expressed milk at discharge or at the time of the first outpatient appointment. No infants received breast milk exclusively.

### Length of Stay and Discharge Coordination

Average length of stay was nine (5-52) days, however, when excluding two premature infants, length of stay decreased to seven days. Discharge to COVID-19 negative family members outside the primary residence occurred in 60% of cases. In 40% of cases, discharge was arranged with fathers who were negative, mothers who were >10 days from their positive test results, or COVID-19 positive parents who could not identify uninfected family members.

## Discussion

We describe infection control measures that resulted in no vertical or horizontal transmission of SARS-CoV-2. Other centers reporting no perinatal transmission adopted similar infection control measures.(17) While measures may decrease transmission, they may also interrupt exchange of maternal flora with newborns. We also report secondary outcomes associated with our policies, including increased length of hospital stay, decreased breastmilk use, and discharge to households outside principal residences.

While C-section rates were higher in our cohort when compared with institutional (29% in 2019) and national rates (31.9% in 2018), our sample size limits interpretation of this result.(18) Maternal complications from COVID-19 may contribute to C-sections. However, in our study, this was the case in only one patient. Other studies of COVID-19 and pregnancy outcomes report an increase in C-sections due to fetal distress even in asymptomatic mothers, which may impact the neonatal microbiome.(17, 19)

Vaginal deliveries represent one of the earliest exposures of newborns to microbes.(20, 21) These pioneer bacteria initiate intestinal eubiosis and impart optimal health outcomes, including decreased incidence of otitis media, atopic disease, and obesity.(22, 23) Without exposure to the vaginal microbiome, the microbiota of infants born via C-sections shifts towards colonization with flora from maternal skin, hospital staff, and hospital environment.(24) Further research is needed to understand factors contributing to an increased incidence of C-sections, particularly in asymptomatic COVID-19 positive women.(25)

In a minority of patients, delayed cord clamping and skin-to-skin care occurred despite institutional policies that advised against such, reflecting the challenges of implementing deviations from routine delivery room care, particularly for term infants. Revised AAP guidelines no longer recommend against delayed cord clamping or skin-to-skin care and our institutional policies have been updated to mirror these changes. Newborns in our cohort were isolated post-partum and then exposed to households outside their parents. New environmental exposures could alter intestinal bacterial colonization early in life. We were not able to evaluate the gut microbiome in our cohort but could be incorporated into future research. Over the first year, newborn microbiota is influenced by mothers’ microbiome and evolves to have a similar composition.(15) Interruptions in this microbial exchange may have unintended consequences later in life, including inflammatory bowel disease, atopic disorders, and obesity.

It is unclear whether separation of infants from mothers had unintended consequences on maternal milk use. Consistent with AAP recommendations, we provided lactation support to encourage expression and storage of milk.(26) However, given ambiguity about COVID-19 transmission through breastmilk and our unit’s milk cataloging system, we restricted maternal breastmilk use in the NICU, including breastfeeding. Data on breastmilk transmission is emerging and a recent study reports no detection of SARS-COV-2 in breast milk from nine infected mothers.(27) However, in one pre-print report, SARS-COV-2 was detected in breastmilk from a symptomatic mother whose infant also subsequently tested positive.(28) Benefits of breastmilk are well understood and implications of not using breastmilk are significant, particularly as it relates to the neonatal microbiome.

In conclusion, while adopting approaches to minimize viral transmission, we and other institutions may have affected various aspects of newborn health, including assembly of the neonatal microbiome. With new information on COVID-19 perinatal transmission, the AAP has revised recommendations, including the safety of delayed cord clamping, mothers and infants rooming together, and breastfeeding.(29) As of November, we revised our policies to allow COVID-19 positive mothers and their offspring to room together without NICU admission. We continue to test infants after delivery and track postnatal outcomes. Meanwhile, a better understanding of the gut-lung axis dysbiosis in COVID-19 is necessary to elucidate potential microbiome-virome cross-talk in utero and postnatally, impact of COVID-19 infection control policies on the neonatal gut microbiome, and ways to safeguard pediatric health outcomes during the pandemic.(30, 31)

## Data Availability

All data pertaining to this manuscript are stored on Box.com through a University Box Health Data Folder, which is a HIPAA compliant tool for secure storage of Protected Health Information.

## Author contributions

JRK, JS, and DMP conceptualized and drafted the article. JRK, DF, JH, and ZK acquired and analyzed data. JRK and JZ developed figures. All authors revised the article and gave final approval for its content.

## Acknowledgements

We would like to acknowledge NIDDK/National Institutes of Health grants NIDDKR01 DK105118 and R01DK114126 (JS).

